# Prognostic Factors for Mortality in Patients Infected with Hantavirus: A Systematic Review with GRADE Certainty Assessment

**DOI:** 10.1101/2024.05.20.24307524

**Authors:** Fernando Tortosa, Fernando Perré, Ignacio Neumann, Martín Alberto Ragusa, Lucia Lossetti, Germán Guaresti, Ariel Izcovich

**Author notes:** Corresponding author: Fernando Tortosa Email address Postal address: Moreno 601, Bariloche, Rio Negro, Argentina.

## Abstract

**Introduction:** One of the challenges in managing patients with hantavirus infection is accurately identifying individuals who are at risk of developing severe disease. Prompt identification of these patients can facilitate critical decisions, such as early referral to an intensive care unit. The identified prognostic factors could be incorporated into predictive models to enhance the management of hantavirus infection.

**Objective:** To identify and evaluate prognostic factors associated with mortality in hantavirus infection, providing a basis for a risk assessment model for hantavirus mortality

**Methods:** We conducted a systematic review following the ‘Preferred Reporting Items for Systematic Reviews and Meta-Analyses’ (PRISMA) guidelines. We conducted a comprehensive search in PubMed/MEDLINE, Cochrane Central Register of Controlled Trials (CENTRAL), and Embase from their inception to January 2024. Furthermore, we included studies evaluating individual prognostic factors or risk assessment models of hantavirus infections, with no restrictions on study design, publication status, or language. When feasible, we conducted meta-analyses for prognostic factors using the inverse variance-based method with random effect model. We assessed the certainty of the evidence using the GRADE approach,

**Results:** We included 30 studies with a total of 92,183 participants. We identified the following key prognostic factors which predicted and increased mortality and disease severity: over 15 years, female gender, elevated creatinine levels (>1.4 mg/dL), increased hematocrit (>42%), and presence of infiltrates on chest radiographs.

**Discussion:** Our systematic review not only sheds light on the pivotal prognostic factors for hantavirus infection but also sets the stage for the development of comprehensive management strategies that are informed by robust empirical evidence. These strategies, underpinned by predictive modeling and regional customization, can significantly enhance outcomes for individuals at risk of severe hantavirus disease, aligning with global health objectives aimed at zoonotic disease control and prevention.

**PROSPERO Registration Number:** CRD42021225823

## Introduction

Hantaviruses, members of the Bunyaviridae family, are single-stranded RNA viruses characterized by their spherical shape, typically measuring 80-100 nm in size.[1]

Unlike other viruses in the Bunyaviridae family, hantaviruses do not rely on arthropods as vectors; instead, they are primarily hosted by rodents and certain small mammals. Each hantavirus genotype tends to associate with specific rodent species. These rodents typically maintain chronic infections with high rates of viral replication, often remaining asymptomatic. Rodent populations fluctuate based on environmental factors such as climate and food availability, with increases in rodent density sometimes correlating with an uptick in human cases. [2–3]

Human transmission primarily occurs through inhalation of infected rodent excreta, occasionally through bites. Person-to-person transmission has been documented for the Andes virus in the Patagonian region of Argentina and Chile.[4]

Prognostic factors, whether used alone or combined in risk assessment models, provide a means of stratifying patients with hantavirus infections based on their risk of developing severe disease or mortality. This stratification can inform optimized treatment strategies and resource allocations. Early identification of patients at risk of deterioration, progression to severe forms of the disease (hantavirus cardiopulmonary syndrome), or higher mortality rates enables prompt initiation of treatment, monitoring, and appropriate support, including timely referrals to specialized intensive care units experienced in managing severe Andes hantavirus infections and venoarterial extracorporeal membrane oxygenation. [5]

While there is consensus on certaincertain prognostic factors, such as low platelet count, the predictive capabilities of many potential prognostic indicators remain uncertain and require further assessment. [6–7]

This systematic review aims to provide a comprehensive summary of the available evidence on prognostic factors for severity and mortality, with the goal of informing clinical decision-making in the management of patients infected with hantavirus and therefore inform a risk assessment model for hantavirus mortality.

## Methods

Our review adheres to the ‘Preferred Reporting Items for Systematic Reviews and Meta-Analyses’ (PRISMA) guidelines [10]. Additionally, we utilized the CHARM-PF checklist for item extraction from primary prognostic factor studies [11]. We published [8] and registered the protocol in PROSPERO (Registration number: CRD42021225823).

### Search strategies

We performed highly sensitive searches in PubMed/MEDLINE, the Cochrane Central Register of Controlled Trials (CENTRAL), and Embase from their inception to January 2, 2024. No restrictions regarding study design, publication status, or language were applied. Detailed search strategies:

## Search strategy

### Search strategy for Pubmed/MEDLINE

#### Last search date: 02.01.2024

#1 hantavirus*

#2 “hantavirus pulmonary syndrome”

#3 ANDV

#4 SNV

#5 “New York virus”

#6 “Black Creek Canal virus”

#7 “Bayou virus”

#8 “White Water Arroyo virus”

#9 “El Moro Canyon virus”

#10 Hantaan virus

#11 Seoul virus

#12 Puumala virus

#13 Dobrava-Belgrade virus

#14 Saaremaa virus

#15 Belgrade virus

#16 Thottapalayam virus

#17 Khabarovsk virus

#18 Amur/Soochong virus

#19 Marah virus

#20 Sangassou virus

#21 Monica virus

#22 Hantaan orthohantavirus

#23 Prospect Hill orthohantavirus

#24 Tula orthohantavirus

#25 Timbo virus

#26 Choclo orthohantavirus

#27 Laguna Negra orthohantavirus

#28 Juquitiba orthohantavirus

#29 Lechiguanas orthohantavirus

#30 #1 OR #2 OR #3 OR #4 OR #5 OR #6 OR #7 OR #8 OR #9 OR #10 OR #11 OR #12 OR #13 OR #14 OR #15 OR #16 OR #17 OR #18 OR #19 OR #20 OR #21 OR #22 OR #23 OR #24 OR #25 OR #26 OR #27 OR #28 OR #29

#31 incidence[MeSH:noexp]

#32 mortality[MeSH Terms]

#33 follow up studies[MeSH:noexp]

#34 prognos*[Text Word]

#35 predict*[Text Word]

#36 course*[Text Word]

#37 #31 OR #32 OR #33 OR #34 OR #35 OR #36

#38 #30 AND #37 AND #38

### Search strategy for EMBASE (Elsevier)

#### Last search date: 02.01.2024

#1. hantavirus*

#2. “hantavirus pulmonary syndrome”

#3. ANDV

#4. SNV

#5. “New York virus”

#6. “Black Creek Canal virus”

#7. “Bayou virus”

#8. “White Water Arroyo virus”

#9. “El Moro Canyon virus”

#10. Hantaan virus

#11. Seoul virus

#12. Puumala virus

#13. Dobrava-Belgrade virus

#14. Saaremaa virus

#15. Belgrade virus

#16. Thottapalayam virus

#17. Khabarovsk virus

#18. “Amur/Soochong virus”

#19. Marah virus

#20. Sangassou virus

#21. Monica virus

#22. “Hantaan orthohantavirus”

#23. “Prospect Hill orthohantavirus”

#24. “Tula orthohantavirus”

#25. “Timbo virus”

#26. “Choclo orthohantavirus”

#27. “Laguna Negra orthohantavirus”

#28. “Juquitiba orthohantavirus”

#29. “Lechiguanas orthohantavirus”

#30. #1 OR #2 OR #3 OR #4 OR #5 OR #6 OR #7 OR #8 OR #9 OR #10 OR #11 OR #12 OR #13 OR #14 OR #15 OR #16 OR #17 OR #18 OR #19 OR #20 OR #21 OR #22 OR #23 OR #24 OR #25 OR #26 OR #27 OR #28 OR #29

#31. ‘incidence’/de

#32. ‘mortality’/exp

#33. ‘follow up’/de

#34. ‘prognosis’/de

#35. ‘predict’/de

#36. ‘course’/de

#37. #31 OR #32 OR #33 OR #34 OR #35 OR #36

#38. #30 AND #37 AND #38

### Search strategy for CENTRAL (The Cochrane Library)

#### Last search date: 02.01.2024

#1. hantavirus*:ti,ab,kw

#2. “hantavirus pulmonary syndrome”:ti,ab,kw

#3. ANDV:ti,ab,kw

#4. SNV:ti,ab,kw

#5. “New York virus”:ti,ab,kw

#6. “Black Creek Canal virus”:ti,ab,kw

#7. “Bayou virus”:ti,ab,kw

#8. “White Water Arroyo virus”:ti,ab,kw

#9. “El Moro Canyon virus”:ti,ab,kw

#10. Hantaan virus:ti,ab,kw

#11. Seoul virus:ti,ab,kw

#12. Puumala virus:ti,ab,kw

#13. Dobrava-Belgrade virus:ti,ab,kw

#14. Saaremaa virus:ti,ab,kw

#15. Belgrade virus:ti,ab,kw

#16. Thottapalayam virus:ti,ab,kw

#17. Khabarovsk virus:ti,ab,kw

#18. Amur/Soochong virus:ti,ab,kw

#19. Marah virus:ti,ab,kw

#20. Sangassou virus:ti,ab,kw

#21. Monica virus:ti,ab,kw

#22. Hantaan orthohantavirus:ti,ab,kw

#23. Prospect Hill orthohantavirus:ti,ab,kw

#24. Tula orthohantavirus:ti,ab,kw

#25. Timbo virus:ti,ab,kw

#26. Choclo orthohantavirus:ti,ab,kw

#27. Laguna Negra orthohantavirus:ti,ab,kw

#28. Juquitiba orthohantavirus:ti,ab,kw

#29. Lechiguanas orthohantavirus:ti,ab,kw

#30. #1 OR #2 OR #3 OR #4 OR #5 OR #6 OR #7 OR #8 OR #9 OR #10 OR #11 OR #12 OR #13 OR #14 OR #15 OR #16 OR #17 OR #18 OR #19 OR #20 OR #21 OR #22 OR #23 OR #24 OR #25 OR #26 OR #27 OR #28 OR #29

#31. MeSH descriptor: [Incidence] this term only

#32. MeSH descriptor: [Mortality] explode all trees

#33. MeSH descriptor: [Follow-Up Studies] this term only

#34. MeSH descriptor: [Prognosis] this term only

#35. MeSH descriptor: [Predictive Value of Tests] this term only

#36. MeSH descriptor: [Disease Progression] this term only

#37. #31 OR #32 OR #33 OR #34 OR #35 OR #36

#38. #30 AND #37 AND #38

We also reviewed the reference lists of each included study and conducted cross-referencing in Google Scholar using each included study as the index reference.

### Study selection

Four reviewers worked independently and in duplicate to perform study selection, which involved screening titles and abstracts as well as potentially eligible full-text articles. Disagreements were resolved through discussion. We included studies that examined individual prognostic factors or risk assessment models based on the typologies of prognosis proposed by Iorio and colleagues [9] and the PROGnosis RESearch Strategy (PROGRESS) Group framework [10], without applying any restrictions based on analytical methods, such as performing multivariable analysis.

### Outcomes

We selected mortality as the only outcome due to its direct clinical relevance in assessing the impact of hantavirus infection. This choice aligns with the need to identify prognostic factors that can predict critical outcomes, facilitating timely interventions for patients at risk of severe consequences.

### Data extraction

For each eligible study, two pairs of reviewers independently abstracted the following information: the year of publication, country, and time period in which the study was conducted for study characteristics; sample size, study context, and other demographic details for population characteristics; details on the prognostic factors examined, their definitions, and outcome measures for description of prognostic factors and outcomes; and measures of association or crude event rates for each candidate prognostic factor and reported outcomes for study results.

### Risk of Bias Assessment

Two reviewers independently assessed the risk of bias of individual included studies using the Quality in Prognosis Studies (QUIPS) tool for prognostic factor studies [11]. We examined population characteristics, attrition, prognostic factors, outcome measurement, and potential residual confounding. Discrepancies were resolved through consensus.

### Data Synthesis and Analysis

We presented the findings of individual prognostic factors both in tabular and narrative formats. To enhance comparability and accuracy, we standardized the units of measurement for each prognostic factor and ensured consistency in the direction of predictors. [12] Whenever feasible, we conducted meta-analyses for prognostic factors and their association with the selected outcomes. To generate an overall measure of association, we utilized the generic inverse variance method. We employed random-effects models based on the DerSimonian-Laird method, using the metafor package in R software [13]

For each candidate prognostic factor, we present the measure of association as odds ratios (OR) along with their corresponding 95% confidence intervals (CIs). In studies that reported the measure of association as a hazard ratio (HR) or risk ratio (RR), we converted them to ORs using the outcome prevalence reported in the studies. [14–15]. When measures of association were not provided for dichotomous variables, we used the crude event rate to calculate ORs. Information on continuous variables without measures of association was excluded. Additionally, we calculated absolute risk differences (RDs) attributable to each individual candidate prognostic factor by applying the ORs to estimated baseline risks (see “Baseline risks” below). In cases where the same candidate prognostic factor was assessed in multiple ways (e.g., dichotomous and continuous), we prioritized the measure for which we found a higher certainty of evidence according to the GRADE approach.

### Assessment of Certainty of Evidence

Assessment of the certainty of evidence was conducted for each candidate prognostic factor and outcome using the GRADE approach [16]. This method evaluates several domains, including risk of bias, indirectness, inconsistency, imprecision, and publication bias. Summary of findings tables were generated, and the certainty of evidence was categorized as high, moderate, low, or very low, depending on the evaluation of each domain [16]. Further details on the assessment of certainty of evidence are provided in S1 File of Supplementary methods.

#### Risk of bias

We used the Quality in Prognosis Studies tool (QUIPS) for prognostic factor studies.^11^ To be rated as low risk of bias studies needed to be prospective, have appropriately assessed prognostic factors (measured at baseline) and outcomes and analyzed the information by considering at least age, one comorbidity and one parameter of disease severity as potential confounders. To be rated as moderate risk of bias studies needed to have appropriately assessed prognostic factors and outcomes and analyzed the information by considering at least one of the pre-defined core set of variables: age, one comorbidity or one parameter of disease severity as potential confounders. The remaining studies were categorized as high risk of bias. RoB was assessed on a study basis but the domain related to considering potential confounder was also assessed on a prognostic factor basis as some studies provided adjusted estimates for some but not all prognostic factors. For the primary analysis we downgraded the certainty of the evidence for risk of bias when no studies with moderate or low risk of bias providing adjusted estimates were available or when subgroup analysis showed inconsistency between moderate/low risk of bias adjusted estimates and high risk of bias studies and the overall pooled estimate of effects was used. We also performed a sensitivity analysis in which we downgraded for risk of bias only when adjusted estimates were not available or when subgroup analysis showed inconsistency between adjusted and unadjusted estimates and the overall pooled estimate was used.

#### Inconsistency

We used visual inspection of the forest plots and the I2 statistic to assess inconsistency. In doing so we considered the variability in point estimates and confidence interval overlap in relation to the thresholds set (see contextualization).

#### Imprecision

We rated down for imprecision when the 95%CI of the pooled estimates crossed the thresholds set (see contextualization). Additionally, we rated down for imprecision if the number of events was less than 200 as we assumed that the optimal information size was not met.^s1^

#### Selective reporting bias

In cases when most of the weight of the pooled estimates were provided by studies in which a multivariable analysis was performed but no adjusted estimates were provided for that particular variable, we considered rating down for selective reporting.

#### Publication bias

Given the nature of our research question (no interventions involved) and the facilities for reporting research results in this specific context (most of identified studies were published as preprint at the moment we performed the search). We assumed that publication bias was not a major issue and did not explore it while addressing certainty of the evidence.

### Result interpretation

To facilitate the interpretation and clinical application of identified prognostic variables, our research team set an arbitrary threshold of a 2% absolute risk increase or decrease, defining a clinically significant escalation in mortality risk. This threshold was established to differentiate between negligible and meaningful changes in risk, aiding clinicians and researchers in assessing the practical importance of each prognostic factor. This approach allows for a clearer understanding of when changes in prognostic indicators become significant enough to influence clinical decisions and public health interventions.

### Baseline risks

For the assessment of baseline risks associated with various prognostic factors, we calculated these risks as percentages based on the prevalence of each factor and their corresponding estimates of association.[28] In instances where specific data on the prevalence of these prognostic factors were not available within the included studies, we utilized described average baseline risks from the literature to estimate these values. This approach ensured a comprehensive evaluation of the potential impact of each prognostic factor on patient outcomes, particularly in informing the risk difference calculations necessary for our analyses.

### Additional analysis

We conducted predetermined sensitivity analyses to determine if risk of bias and adjustment of study parameters could act as effect modifiers. These analyses were specifically tailored to evaluate the stability of our findings under various scenarios and configurations.

We also conducted subgroup analyses to differentiate the impact of various hantavirus subtypes, particularly focusing on the Andes virus compared to other viruses. This analytical approach was employed due to the distinct epidemiological and clinical features of the Andes virus, including its unique potential for person-to-person transmission and a potentially higher mortality rate, which is not commonly observed in other hantavirus subtypes. Additionally, the presence of outbreaks with high mortality rates in the region further underscores the critical nature of studying this particular subtype. In cases where a potential effect modification was identified, we assessed the credibility of these findings using the criteria defined in the ICEMAN tool. [49]

## Results

We initially identified 3,382 records through various databases. After removing 1,565 duplicates, we reviewed 1,817 articles by title and abstract. After full-text assessment, we included 30 primary studied in the review, as detailed in Figure 1.

**Figure 1.**
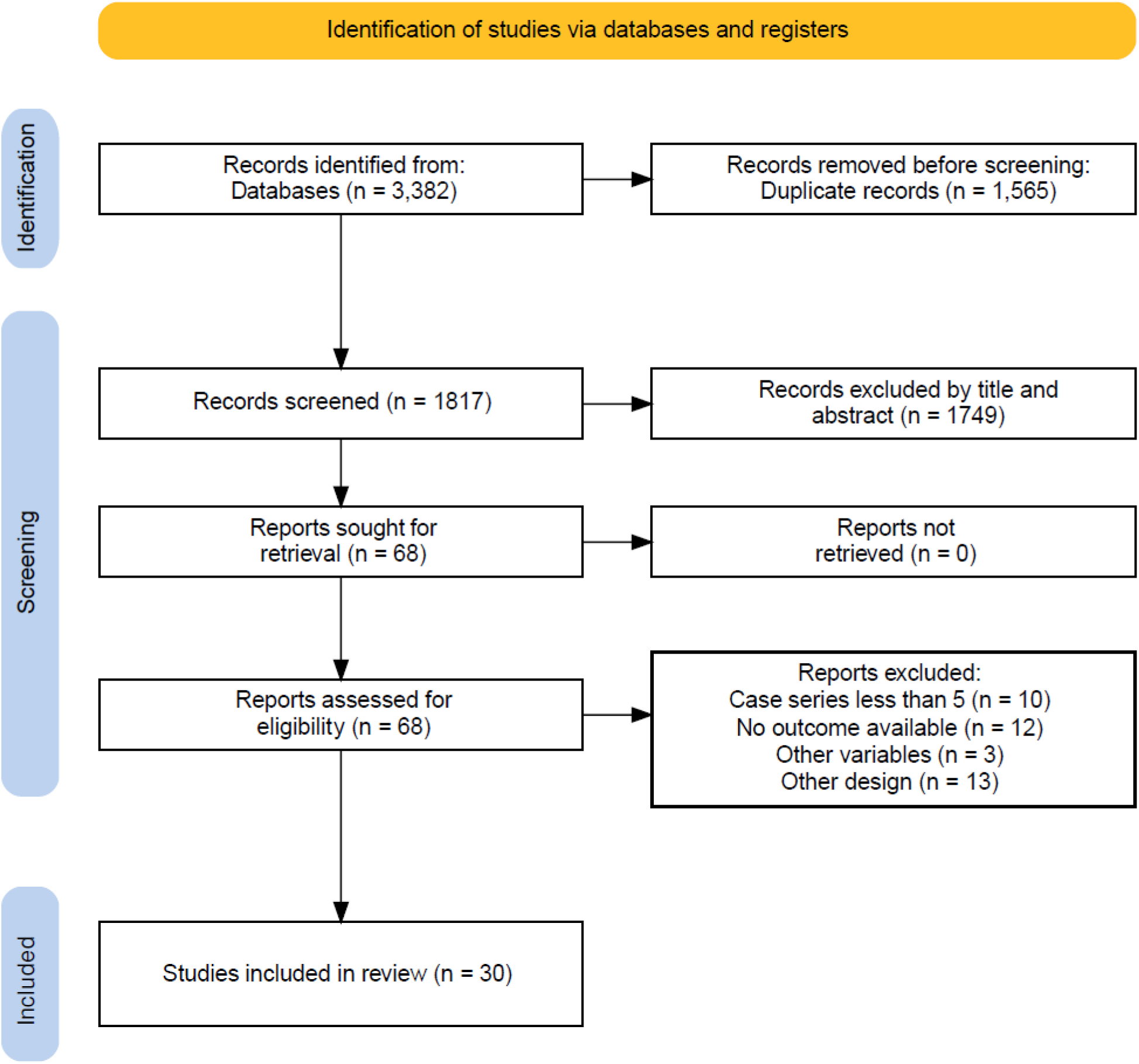
**Identification of studies via databases and registers**

### Description of included studies

The following variables were assessed as potential prognostic factors in the 30 studies included in our review. The age over 40 years was evaluated in 9 studies with 83,971 participants (the studies primarily focused on age groups below 19, while also including broader categorizations that spanned from under 10 to over 60 years, with specific age thresholds at 40 and 50 years in several studies); female gender in 18 studies with 88,435 patients; duration of symptoms over 5 days in 1 study with 123 participants; and hematocrit over 42 in 5 studies with 737 participants. Additionally, the presence of proteinuria in urine was considered in 2 studies with 170 participants, signs of bleeding in 4 studies with 1,416 participants, infiltrates on chest radiograph in 6 studies with 1,998 participants, and a platelet count less than 100,000/mm³ in 4 studies with 886 participants. The increase in AST by 3 times was analyzed in 2 studies with 479 participants, the increase in ALT by 3 times in 1 study with 24 participants, the presence of vomiting as a predominant symptom in 1 study with 144 participants, and the presence of headache as a predominant symptom in 2 studies with 523 participants. Finally, leukocytosis greater than 12,000/mm³ was analyzed in 5 studies that covered 1,864 participants. Details on the studies excluded can be found in the For more information about included studies see Table 1..

**Table 1.** Characteristics of included studies.

| First Author | Study year | Geographic location | N | Age (mean) | % SPH at start | N HPS progression | N deaths | Diagnosis | Viral serotype |
| --- | --- | --- | --- | --- | --- | --- | --- | --- | --- |
| Wernly | 2011 | USA | 51 | 39.6 years | 100% | NA | 17 | IgM/IgG | SNV |
| Warner | 2020 | Canada (1989-2019) | 143 | (median 42, range 7–76 years of age) | 69% | NR | 34 | ELISA IgM and RT-PCR | SNV |
| Vial | 2019 | Chile | 139 | 37 years, range 10-77 years | NR | 46% |  | ELISAs, o RT-qPCR para ANDV | ANDV |
| Tortosa | 2022 | Argentina (1996-2022) | 123 | 35 years (DS 17.1) | 28.40% | 70.40% | 50 | IgM en dos muestras separadas por 5 días y RT-PCR | ANDV |
| Riquelme | 2015 | Chile | 80 | 35.7 ± 16 | 100% | NA | 24 | PCR-RT | ANDV |
| Ramos | 2001 | USA | 11 | 14 (10-16) | NR | 45.45% | 2 | IgM/IgG | SNV |
| Pantozzi | 2011 | Argentina | 291 | median 28 years (range 15 to 49 years) | NR | 42.10% | 79 | ELISA IgM and RT-PCR<sup>a</sup> | ANDV |
| Oliveira | 2015 | Brazil | 251 | median 34.56 +/- 13.38 years (rango 0-73 years) | 100 | NA | 75 | ELISA IgM and RT-PCR | JUQV |
| Menezes | 2016 | Brazil | 73 | 34.9 (SD +/- 13.8 years) | 57.50% | NA | 42 | ELISA para IgM | Araraquara and Juquitiba |
| Maleki | 2019 | Argentina | 93 | 36 ± 16 years | 100 | NA | 34 | ELISA IgM and RT-PCR | ANDV |
| MacNeil | 2011 | USA | 510 | median 38 years (20-50 years) | 35% | NA | 178 | HPS detection of hantavirus-specific IgG y PCR, | SNV, Bayou Virus, New York Virus, Monogahela virus, Black Creek Canal virus |
| López R | 2019 | Chile | 40 | median 35 years (23-46 years) | 0% | 14 | 4 | ELISA IgM and RT-PCR | ANDV |
| López R | 2021 | Chile | 175 | median 35 ayears (22-47 years) | 100% | NA | 37 | ELISA IgM and RT-PCR | ANDV |
| Limongi | 2007 | Brazil | 23 | median 23 years (29+/- 13.6 years) | 100% | NA | 5 | ELISA IgM and RT-PCR | Araraquara |
| Korva | 2019 | United States | 510 | median 38 years (range 20-50) | 100% | NA | 132 | ELISA IgM | Sin Nombre virus |
| Klein | 2011 | China (1996-2009) | 80671 | 95% more than 20 years | NA | NA | 945 | ELISA IgM and RT-PCR | PUUV |
| Iglesias | 2015 | Argentina | 86 | 79% more than 20 years | NR | NR | 22 | ELISA IgM and RT-PCR | ANDV |
| Fonseca | 2020 | Brazil (2007-2015) | 1004 | 98% more than 10 years | NR | NR | 410 | NR | NR |
| Du | 2014 | India | 75 | 48.76 ± 13.59 years | NR | NR | 29 | ELISA IgM-iGg | HTNV |
| Du (b) | 2014 | China (2008-2012) | 356 | 47.88±14.78 years | 35% | 160 | 29 | ELISA IgM-iGg | Hantaan virus (HTNV) and Seoul virus (SEOV) |
| Du | 2021 | China (2012-2014) | 106 | 41.85 ± 15.35 years | 58% | NR | 61 | ELISA IgM-iGg | HTNV |
| da Rosa Elkhoury | 2012 | Brazil (1993-2006) | 855 | 33 years (20-39) | 100 | NA | 336 | ELISA IgM-iGg | Juquitiba, Araraquara, Castelo dos Sonhos, Anajatuba and Rio Mearim |
| Castillo | 2001 | Chile (1997-1999) | 13 | 30 years (range, 19 to 45 years) | 100 | NA | 7 | ELISA IgM-iGg | Andes virus (ANDV) |
| Campos | 2009 | Brazil (1998 and 2007) | 70 | 35,8±11,7 years | 61.43% | 29 | 38 | ELISA IgM and RT-PCR | vírus Sin Nombre (SNV); Andes virus (ANDV) |
| Arita | 2019 | Brazil (1992-2016) | 280 | NR | NR | NR | 107 | NR | vírus Araucária e Jaborá |
| He | 2023 | China (2011-2019) | 15 | 42.9±14.6 | NA | NA | 317 | NR | Hantaan virus (HTNV) and Dobrava (DBV), Pumaala (PUUV) |
| Ferres | 2010 | Chile (1997-2007) | 24 | Under 13 years | 62.50% | NR | 7 | ELISA IgM and RT-PCR | Andes virus (ANDV) |
| Santana | 2006 | Brazil (1993-2004) | 27 | NR | 100% | NR | 13 | ELISA IgM and RT-PCR | vírus Sin Nombre (SNV) |
| Korva | 2019 | Slovenia (2000-2014) | 170 | NR | NA | NA | 17 | ELISA IgM and RT-PCR | Crimea Congo, Dobrava (DBV), Pumaala (PUUV) |
| Hjertqvist | 2010 | Sweden (1997-2007) | 5282 | 49.3 ± 16.7 years for men and 50.7 ± 16.5 years for women | NA | NA | 21 | ELISA IgM and RT-PCR | Hantaan virus (HTNV) and Dobrava (DBV), Pumaala (PUUV) |

### Risk of bias of included studies

Of the 30 included studies, only 5 were classified as having a low overall risk of bias, as detailed in the S2 Table of Risk of Bias of Included Studies. We identified several key methodological limitations across the body of research that predominantly affect the reliability and validity of outcomes: Most studies were retrospective and used administrative databases. Also, most studies failed to adjust for known confounding factors, such as the severity of the disease at the onset or other relevant clinical variables.

### Prognostic Factors for Mortality in Hantavirus Infection

Table 2 provides a summary of findings for all the identified candidate prognostic factors. We found high or moderate certainty that the following factors may increase the risk of death in hantavirus infection (S1-20 Figs).

**Table 2.** Summary of findings table.

| Prognostic Factor | N studies | N participants | Pooled OR (95% CI) | Risk without Prognostic Factor (%) | Risk Difference with PF (95% CI) | Overall Certainty | Interpretation |
| --- | --- | --- | --- | --- | --- | --- | --- |
| Age (>40 years) | 9 | 83,971 | 1.95 (1.32, 2.88) | 24.0 | 11.2% more (from 5.5% more to 15.2% more) | Moderate<sup>e</sup><br>⊕⊕⊕⊖ | Age more than 15 years probably a prognostic factor for death |
| Female Gender | 18 | 88,435 | 1.35 (1.23, 1.48) | 32.3 | 6.7% more (from 4.6% more to 8.7% more) | Moderate<sup>e</sup><br>⊕⊕⊕⊖ | Female sex probably a prognostic factor for death |
| DIS > 5 days | 1 | 123 | 1.46 (0.70, 3.02) | 29.2 | 7.8% more (from 8.5% less to 18.8% more) | Very Low<sup>c</sup><br>⊕⊖⊖⊖ | Uncertain whether DIS for more than 5 days is a prognostic factor |
| Hematocrit > 42 | 5 | 737 | 1.8 (1.32, 2.46) | 28.1 | 11.9% more (from 6.0% more to 17.0% more) | Moderate<sup>e</sup><br>⊕⊕⊕⊖ | Hematocrit > 42 probably a prognostic factor for death |
| Atypical Lymphocytes | 1 | 182 | 1.22 (0.66, 2.26) | 32.6 | 4.4% more (from 9.5% less to 16.3% more) | Very Low<sup>c</sup><br>⊕⊖⊖⊖ | Uncertain whether presence of atypical lymphocytes is a prognostic factor |
| Increased Creatinine (>1.4 g/dl) | 7 | 2,126 | 1.58 (1.36, 1.84) | 30.1 | 9.9% more (from 6.7% more to 12.9% more) | Moderate<sup>e</sup><br>⊕⊕⊕⊖ | Increased creatinine (>1.4 g/dl) probably a prognostic factor for death |
| Proteinuria in Urine | 2 | 170 | 2.29 (0.48, 10.92) | 23.1 | 13.9% more (from 20.6% less to 24.0% more) | Very Low<sup>c</sup><br>⊕⊖⊖⊖ | Uncertain whether presence of proteinuria in urine is a prognostic factor |
| Signs of Bleeding | 4 | 1,416 | 1.98 (1.14, 3.43) | 31.0 | 15.0% more (from 2.9% more to 25.2% more) | Low<sup>a-d</sup><br>⊕⊕⊖⊖ | Presence of bleeding could be a prognostic factor |
| Infiltrates on Chest Radiograph | 6 | 1,998 | 2.72 (1.36, 5.46) | 24.4 | 17.6% more (from 6.5% more to 24.4% more) | Moderate<sup>e</sup><br>⊕⊕⊕⊖ | Infiltrates on chest radiograph probably a prognostic factor for death |
| Platelet Count < 100,000/mm<sup>3</sup> | 4 | 886 | 1.37 (0.69, 2.73) | 30.4 | 6.6% more (from 8.9% less to 17.3% more) | Very Low<sup>c</sup><br>⊕⊖⊖⊖ | Uncertain whether presence of platelet count less than 100,000/mm<sup>3</sup> is a prognostic factor |
| Elevated AST x 3 | 2 | 479 | 3.91 (0.65, 23.58) | 30.4 | 29.6% more (from 9.2% less to 48.8% more) | Very Low<sup>c</sup><br>⊕⊖⊖⊖ | Uncertain whether presence of elevated AST x 3 is a prognostic factor |
| Elevated ALT x 3 | 1 | 24 | 1.15 (0.22, 6.02) | 33.8 | 3.2% more (-26.8% less to 34.8% more) | Very Low<sup>c</sup><br>⊕⊖⊖⊖ | Uncertain whether presence of elevated ALT x 3 is a prognostic factor |
| Presence of Vomiting | 1 | 144 | 0.68 (0.33, 1.38) | 36.9 | 8.9% less (from 24.9% less to 7.0% more) | Very Low<sup>c</sup><br>⊕⊖⊖⊖ | Uncertain whether presence of vomiting is a prognostic factor |
| Presence of Headache | 2 | 523 | 0.34 (0.03, 3.76) | 51.0 | 20.0% less (from 62.1% less to 3.9% more) | Very Low<sup>c</sup><br>⊕⊖⊖⊖ | Uncertain whether presence of headache is a prognostic factor |

| Certainty |  | Explanation |
| --- | --- | --- |
| High | ⊕⊕⊕⊕ | We are very confident about this prognostic factor |
| Moderate | ⊕⊕⊕⊖ | We are somewhat confident about this prognostic factor |
| Low | ⊕⊕⊖⊖ | We are not very confident about this prognostic factor |
| Very Low | ⊕⊖⊖⊖ | We are uncertain about this prognostic factor |

**Notes:**
- a. We downgraded one level due to imprecision. - b. We downgraded two levels due to severe imprecision. - c. We downgraded three levels due to very severe imprecision - d. We downgraded one level due to inconsistency - e. We downgraded one level due to risk of bias.

Symptoms, Vital Signs, and Physical Examination Factors: Age greater than 40 years: OR 1.95, 95% CI 1.32 to 2.88, risk difference (RD) 11.2% (95% CI 5.5% to 15.2%, moderate certainty evidence). Female Gender: OR 1.35, 95% CI 1.23 to 1.48, RD 6.7% (95% CI 4.6% to 8.7%, moderate certainty evidence).

Laboratory Factors (Measured in Blood or Plasma): Increased Creatinine (>1.4 mg/dL): OR 1.58, 95% CI 1.36 to 1.84, with a risk difference of 9.9% (95% CI 6.7% to 12.9%, moderate certainty evidence), Hematocrit > 42%: OR 1.80, 95% CI 1.32 to 2.46, with a risk difference of 11.9% (95% CI 6.0% to 17.0%, moderate certainty evidence).

Radiological Findings: Infiltrates on Chest Radiograph: OR 2.72, 95% CI 1.36 to 5.46, with a risk difference of 17.6% (95% CI 6.5% to 24.4%, moderate certainty evidence). For detailed information on each factor and their impact, please refer to Table 2: Summary of findings table.

### Additional analysis

The predetermined sensitivity analyses conducted, which included testing for risk of bias and adjustments for the parameters of the studies, revealed that the results were robust. We found no substantial evidence indicating that the assessed parameters are effect modifiers.

The only potential prognostic factor for which the type of virus might be an effect modifier is proteinuria (S10 Fig). We found that proteinuria might be associated with increased mortality in Andes virus patients, with an odds ratio (OR) of 5.73 (95% CI: 1.63, 20.14) and a risk difference of 21.4% more (ranging from 9.3% to 25.3%), although the certainty of this evidence is low (S3 Table). Other factors, such as increased creatinine and leukocytosis, showed very low certainty, indicating unclear prognostic significance. However, when analyzing the reliability of this subgroup effect using the criteria described in the ICEMAN tool, the result indicates very low credibility.

## Discussion

We identified several critical factors associated with increased mortality risks in individuals with hantavirus infection. Moderate certainty evidence suggested that age over 40 years, female gender, elevated creatinine levels (>1.4 mg/dL), increased hematocrit (>42%), and the presence of infiltrates on chest radiographs all correlate with higher mortality.

In our analysis, proteinuria emerged as a potential prognostic factor for increased mortality among Andes virus patients. However, the very low credibility of this finding, as assessed using the ICEMAN criteria, advises caution in treating it as a definitive effect modifier. Variability and potential biases in the studies considered may have influenced these results. Other factors, such as increased creatinine levels and leukocytosis associated with the Andes virus subtype, displayed very low certainty and ambiguous prognostic value, emphasizing the difficulties in pinpointing reliable prognostic markers for hantavirus infections.

This systematic review is the first to comprehensively evaluate multiple candidate prognostic factors, assess their impact on mortality, and determine the certainty of these impacts using the GRADE methodology. It addresses a gap left by previous studies, which often provided unadjusted values and were based on very low certainty evidence.

Considering the variables identified with moderate certainty, and the scarcity of prognostic information for hantavirus infection, these factors could be crucial for developing a prognostic scoring system or evaluation method. Such a tool would be invaluable in clinical settings to assess patient mortality risk. Notably, factors like age over 40 years, elevated hematocrit levels, and specific radiological findings could be used to stratify patients effectively into risk categories—deeming older adults with elevated hematocrit high risk, while considering younger patients without radiological infiltrates or significant lab abnormalities low risk.

This provisional assessment tool would significantly aid in guiding clinical decisions and resource allocation until more comprehensive and definitive prognostic methods are developed. Integrating these factors into routine clinical assessments could improve our ability to predict outcomes and tailor interventions to individual patient needs, thus enhancing overall management of hantavirus infection.

Our review also showcases strengths such as its comprehensive scope and rigorous methodology following the GRADE approach, aligning with current standards in the field. By detailing the absolute contributions of various prognostic variables and setting a clear criterion for clinical significance, we enhance the usability of our findings.

However, the review has limitations due to significant variability in the characteristics of the included studies, such as design, patient eligibility criteria, and definitions of prognostic factors. This diversity posed challenges in conducting subgroup analyses to account for these differences, potentially affecting the generalizability and applicability of our findings across various populations or settings. The variability also restricted our ability to conduct more advanced analyses to establish the independence of the identified prognostic factors.

## Data Availability

All relevant data are within the manuscript and its Supporting Information files.

